# Assessment of neuropsychological function in brain tumour treatment: A comparison of traditional neuropsychological assessment with app-based cognitive screening

**DOI:** 10.1101/2020.06.03.20119255

**Authors:** Rafael Romero-Garcia, Mallory Owen, Alexa McDonald, Emma Woodberry, Moataz Assem, Pedro Coelho, Rob C Morris, Stephen J Price, Tom Santarius, John Suckling, Tom Manly, Yaara Erez, Michael G Hart

**Author notes:** Correspondence to: Rafael Romero-Garcia, Herchel Smith Building for Brain and Mind Sciences, Robinson Way, Cambridge CB2 0SZ. These authors contributed equally (Rafael Romero-Garcia, Mallory Owen).

## Abstract

**Background:** Gliomas are typically considered to cause relatively few neurological impairments. However, cognitive difficulties can arise, for example during treatment, with potential detrimental effects on quality of life. Accurate, reproducible, and accessible cognitive assessment is therefore vital in understanding the effects of both tumour and treatments. Our aim is to compare traditional neuropsychological assessment with an app-based cognitive screening tool in patients with glioma before and after surgical resection. Our hypotheses were that cognitive impairments would be apparent, even in a young and high functioning cohort, and that app-based cognitive screening would complement traditional neuropsychological assessment.

**Methods:** Seventeen patients with diffuse gliomas completed a traditional neuropsychological assessment and an app-based touchscreen tablet assessment (OCS-BRIDGE) pre- and post-operatively. The app assessment was also conducted at 3- and 12-month follow-up. Impairment rates, mean performance, and pre- and post-operative changes were compared using standardized Z-scores.

**Results:** Approximately 2-3 hours of traditional assessment indicated an average of 2.88 cognitive impairments per patient, whilst the 30-minute screen indicated 1.18. As might be expected, traditional assessment using multiple items across the difficulty range proved more sensitive than brief screening measures in areas such as memory and attention. However, the capacity of the screening app to capture reaction times enhanced its sensitivity, relative to traditional assessment, in the area of non-verbal function. Where there was overlap between the two assessments, for example digit span tasks, the results were broadly equivalent.

**Conclusions:** Cognitive impairments were common in this sample and app-based screening complemented traditional neuropsychological assessment. Implications for clinical assessment and follow-up are discussed.

## INTRODUCTION

Patients with diffuse glioma can develop impairments in multiple cognitive domains before or after surgery that can have a profound effect on function and quality of life (Ek et al., 2018; Reijneveld et al., 2016). The type of impairment can be influenced by tumour morphology, extent of resection and post-surgical treatment, tumour recurrence, age and concurrent psychological distress (Anderson et al., 1999; Duffau, 2005; Kaleita et al., 2004; Lang et al., 2017; Lawrie et al., 2019; C. A. Meyers & Hess, 2003; Taphoorn & Klein, 2004; Wefel et al., 2011, 2016). However, in any given case, it is difficult to make precise predictions about cognitive outcome due to e.g. individual differences and long-ranging effects of diffuse gliomas on networks throughout the brain (Harris et al., 2014; Hart et al., 2019; Romero-Garcia et al., 2020; van Dellen et al., 2012). Assessment of cognitive function is therefore vital in informing management and in monitoring the long-term effects of tumours and interventions.

Despite recognition of this importance, several issues remain with current diffuse glioma neurocognitive testing. In a traditional cognitive assessment, a neuropsychologist works through a range of in-depth assessments with the patient, writing down the responses then manually scoring and writing a report. The range of tests reflects the fractionation of the cognitive system, that is distinct functions that can be selectively impaired. For example, in the area of memory, a neuropsychologist may apply different tests of verbal and non-verbal memory, recognition memory vs. recall, autobiographical memory, prospective memory, working memory and so on. These tests typically present many items, graded in difficulty, such that the tests are sensitive to individual differences in the general population as well as simply detecting ‘impairment’.

This approach has great strengths but also important limitations. In-depth assessment of an individual can take several hours, in addition to scoring and report writing time. A long assessment may be highly sensitive in the general population but be overly influenced by fatigue, pain, or other factors in clinical groups (Taphoorn & Klein, 2004). There is such a wide range of specialized tests available that there may be little overlap between those used in one centre and another, making comparison of outcomes complex (Ng et al., 2019; Rofes et al., 2017; Satoer et al., 2016). Many tests have strong or unknown practice effects, limiting their use for repeated assessments.

Computerised cognitive assessments offer potential advantages including highly standardized administration, the capacity to measure reaction times with millisecond precision and instant scoring/reporting. In this way, indications of possible impairment in language, memory, orientation, attention, numerical cognition, spatial bias and ability to imitate gestures can be derived in around 10 minutes, helping to inform management in settings where patients may be intolerant of longer assessments (e.g. acute stroke). The advent of the touchscreen tablet added portability and suitability for bedside use, greater hygiene, a more intuitive means of interaction, and further reduced costs. OCS-BRIDGE is a recently developed tablet app that is a hybrid between very brief screening and more in-depth assessment (https://ocs-bridge.com/) (Demeyere et al., 2015; Mancuso et al., 2018).

To date, there are no reports of the use of app-based cognitive assessment in diffuse glioma. Here 17 patients were given a traditional neuropsychological assessment and OCS-BRIDGE assessment before and after surgery. In addition, OCS-BRIDGE was re-administered at 3- and 12-month follow-up. Our interest was whether OCS-BRIDGE offered advantages in terms of ease of administration and efficiency of repeated testing in longitudinal studies, whether its results accorded well with traditional in-depth assessment, and whether aspects of the two approaches may be complementary.

## MATERIALS AND METHODS

### Participants

This study is a single centre prospective cohort design approved by the Cambridge Central Research Ethics Committee (protocol number 16/EE/0151). All procedures contributing to this work comply with the ethical standards of the relevant national and institutional committees on human experimentation and with the Helsinki Declaration of 1975, as revised in 2008. All patients gave written informed consent. Patients deemed to have typical appearances of a diffuse glioma were identified at adult neuro-oncology multidisciplinary team (MDT) meetings at Addenbrooke’s Hospital (Cambridge, UK), and a consultant neurosurgeon directly involved in the study identified potential patients based on the outcome of the MDT discussion. Patients (n=17) were recruited between 2017-2019 and followed up until 2020. Inclusion criteria included: (*i*) participant is willing and able to give informed consent for participation in the study, (*ii*) imaging is evaluated by the MDT and judged to have typical appearances of a diffuse glioma, (*iii*) Stealth MRI is obtained (routine neuronavigation MRI scan performed prior to surgery), (*iv*) World Health Organisation (WHO) performance status 0 or 1, (*v)* age between 18 to 80 years, (*vi*) tumour located in or near eloquent areas of the brain thought to be important for speech and executive functions, and (*vii*) patient undergoing awake surgical resection of a diffuse glioma. This last inclusion criterion was adopted to collect additional intraoperative electrocorticography data that will be reported separately (Erez et al., 2020). Participants were excluded if any of the following applied: (*i*) concomitant anti-cancer therapy, *(ii)* concomitant treatment with steroids, (*iii*) history of previous malignancy (except for adequately treated basal and squamous cell carcinoma or carcinoma in-situ of the skin) within 5 years, and (*iv*) previous severe head injury. See **Table 1** for demographic and clinical characteristics.

**Table 1.** Demographic Information. SFG, Superior Frontal Gyrus; MFG, Middle Frontal Gyrus; IFG, Inferior Frontal Gyrus; ITG, Inferior Temporal Gyrus; MTG, Middle Temporal Gyrus; SMA, Supplementary Motor Area. Ages have been rounded to off for anonymity.

| Patient | Age | Gender | Handedness | Presentation | Hemisphere | Location | Tumour Grade / Pathology |
| --- | --- | --- | --- | --- | --- | --- | --- |
| 1 | 40 | Female | Left | Seizures | Left | Frontal | Grade II Oligodendroglioma |
| 2 | 30 | Male | Right | Seizures | Right | Insula | Grade II Astrocytoma |
| 3 | 30 | Male | Right | Seizures | Left | Temporal / Insula | Grade IV Glioblastoma |
| 4 | 50 | Female | Right | Incidental | Right | Insula | Grade II Oligodengroglioma |
| 5 | 60 | Female | Right | Recurrence | Left | Frontal / SFG / frontal pole | Grade II Oligodendroglioma |
| 6 | 20 | Female | Left | Seizures | Right | Frontal / IFG | Grade I Ganglioglioma |
| 7 | 30 | Male | Right | Seizures | Right | Frontal / SFG & MFG | Grade III Astrocytoma |
| 8 | 30 | Male | Right | Seizures | Right | Frontal / MFG | Grade III Astrocytoma |
| 9 | 50 | Male | Left | Seizures | Left | Temporal / ITG | Grade IV Glioblastoma |
| 10 | 40 | Female | Right | Seizures | Right | Frontal / MFG | Grade II Oligodendroglioma |
| 11 | 30 | Male | Right | Seizures | Left | Frontal / SFG / frontal pole | Grade II Astrocytoma |
| 12 | 30 | Female | Right | Headaches | Left | Temporal / MTG | Grade III Astrocytoma |
| 13 | 30 | Female | Right | Seizures | Left | Superior Temporal Gyrus | Grade I Ganglioglioma |
| 14 | 60 | Female | Right | Seizures | Left | Superior Temporal Gyrus | Grade II Astrocytoma |
| 15 | 30 | Male | Right | Seizures | Left | Superior Temporal Gyrus | Grade III Astrocytoma |
| 16 | 30 | Male | Right | Seizures | Left | SFG/SMA & Pre-central | Grade IV Glioblastoma |
| 17 | 30 | Male | Right | Seizures | Left | Inferior frontal | Grade III Astrocytoma |

### Cognitive Assessment

Cognitive assessment was carried out using two distinct methods at various time points before and after surgery.

#### Traditional neuropsychological assessment

The neuropsychological battery comprised 26 independent measures of cognitive function across eight domains: verbal memory, nonverbal memory, verbal skills, nonverbal skills, attention, executive function, and mood disturbance using a variety of previously validated tests (J. E. Meyers et al., 2013; Quental et al., 2013; Vlaar & Wade, 2003). The battery included elements of both the Weschler Adult Intelligence Scale IV, which has previously been used to assess cognitive functioning in glioma patients (Zhou et al., 2010), and the Brain Injury Rehabilitation Trust Memory and Information Processing Battery, which was specifically designed for patients with neurological injuries (Crawford & Garthwaite, 2007). Testing took approximately 2-3 hours to complete and was administered by a registered neuropsychologist in a clinical setting. A full list of the tests included on both the OCS-BRIDGE and the neuropsychological assessments can be found in **Table S1**.

#### OCS-BRIDGE Assessment

This novel app-based screening tool, administered via touchscreen tablet, consists of 3 parts. The first part is based on the Oxford Cognitive Screen (OCS) and consists of a brief screening of language, orientation, attention, perception, memory, praxis and numeracy skills, based on a paper and pencil measure extensively validated in stroke (Demeyere et al., 2015; Mancuso et al., 2018). The second part of OCS-BRIDGE provides more sensitive measures, adapted from well-established test paradigms, for patients able to tolerate a slightly longer assessment of 25-40 minutes. It includes measures of reaction time, working memory and visual perception. Screening included 6 memory tasks: free verbal memory, overall verbal memory, episodic memory, orientation (spatial memory), forward and backwards digit span (verbal short term and working memory) (van der Hurk & Hodges, 1995; Wilson et al., 2005). Because performance benefits from previous exposure to the tasks (practice effect) tend to be less marked on such measures, they also lend themselves to repeat assessment in longitudinal follow-up. The final part of OCS-BRIDGE consists of the widely used mood measures PHQ-9 and GAD-7 (Kroenke et al., 2001; Spitzer et al., 1999). OCS-BRIDGE was administered pre-operatively, post-operatively prior to discharge, as well as at 3-month and 12-month follow-up, and took between 20-35 minutes to complete.

#### Cognitive impairments quantification

An impairment in either assessment technique was defined by convention as performance two standard deviations below the mean of a reference control population on any particular test or test component. OCS-BRIDGE offers a second category of ‘possibly impaired’ for scores that fall between approximately the 5^th^-10^th^ percentile. The total impairments in each domain were defined as the number of individual tests within that domain on which a participant demonstrated an impairment.

Individual domains from the neuropsychological battery and the OCS-BRIDGE assessment were combined into four generalized functional domains for the purposes of direct comparison: attention, memory, verbal skills, and nonverbal skills (**Table S2)**. The neuropsychological battery also included two tests of executive function that provided four independent measures, as well as two assessments of mood. These were included in the analysis as their own domains.

Neuropsychological and OCS-BRIDGE values were z-scored by subtracting the mean and dividing by the standard deviation derived from normative healthy participants. Normative data for neuropsychological assessments were obtained from participants aged 16 to 89. OCS-BRIDGE reference scores were derived from 268 healthy controls (Mean Age, 51.44, SD Age 19.86). These samples were representative of the general distribution in terms of age, gender and educational level.

Neurological/health conditions that were likely to impact on scores (e.g. stroke, epilepsy, medications, uncorrected hearing loss etc.) were an exclusion criterion. Further details on the normative data can be found in each individual test manual.

### Statistical Analysis

All statistical analysis was carried out in R (R Core Team, 2014) and Matlab R2021b. The Z-score of a given domain was calculated as the median Z-score of all the items belonging to that domain. We additionally calculated the ratio of cognitive deficits for each patient as the number of tasks showing a deficit (performance below threshold) divided by the total number of tasks. The t-value and p-value were calculated using an unpaired t-test between assessments across patients. The number of tasks that contributed to each domain differed and full details are available in **Table S1**.

Although the number of participants in the current study is low for formal analysis, for illustration, unsupervised clustering analysis was carried out using the K-means algorithm. Individual tests that did not show any variation in score between participants were omitted from the clustering analysis, excluding 14/39 OCS-BRIDGE, and 1/28 Neuropsychological, measures. Clustering was performed on values that had been centered to a mean value of 0 with a standard deviation of 1. K-means clustering was done with a k value of 4 based on the within-groups sum of squares. Analysis was performed using the kmeans function in R version 3.6.1. Clusters were visualized by performing a principal components analysis (PCA) using the fviz function in the factoextra package and plotting the individual cognitive tests against the first and second principal components.

## RESULTS

### Demographics and Data Completeness

No significant difference was found in total impairments reported preoperatively between right and left-handed participants (t=0.547, *P*=0.6183), male and female (t=01.175, *P*=0.258), or between right- and left-sided tumours (t=0.845, *P*=0.411). The correlation between age and impairment frequency did not reach statistical significance (*r*=-0.22, *P*=0.39).

All participants completed pre-operative traditional neuropsychology and OCS-BRIDGE assessments. Fourteen participants completed traditional neuropsychology between two and five weeks after surgery, and eight completed postoperative OCS-BRIDGE assessments within 72 hours. Eleven participants completed an OCS-BRIDGE assessment at 3-month follow-up and at 12-month follow-up. Sixteen of seventeen participants had at least one OCS-BRIDGE assessment after surgery (see **Table S3**).

### Neuropsychiatric Function

In this sample, anxiety was at a relatively low level with just 3/17 participants pre-operatively scoring within the mild range on the Beck Anxiety Index (BAI) and 2/17 in the moderate range. There was no association between mood status and frequency of cognitive impairment in this sample (BAI status – total cognitive impairments t=1.2789, *P*=0.22; BDI status – total cognitive impairments t=0.87, *P*=0.4).

### Traditional Neuropsychological Assessment

On traditional neuropsychological assessment, pre-surgically, 79% (14/17) of participants had an impairment in at least one domain, with a mean of 2.88 (SD = 2.47) impairments per participant. Three participants performed above cut-off in all domains. Impairments were observed in the domains of attention (7), verbal memory (7), verbal skills (6), nonverbal memory (4), and executive function (3). Of the 26 cognitive measures in the battery, 16 detected at least one impaired level of performance among the participants whilst 10 returned no impaired scores for any participant.

After surgery, all but one participant had an impairment in at least one domain, with a mean of 4.50 impairments per participant (SD = 3.40). Compared with testing before surgery, the total number of cognitive impairments apparently reduced in 43% (6/14) participants, remained unchanged in 7% (1/14), and increased by 50% (7/14) (**Figure 1**). Of the 7 participants with more impairments, the average increase was 4.14 (SD 3.18). Three domains (verbal memory, nonverbal memory, and verbal skills) demonstrated an increase in impairments after surgery, two domains (nonverbal skills and attention) remained unchanged, and one (executive function) improved. No participants demonstrated an impairment in nonverbal skills at any point.

**Figure 1.**
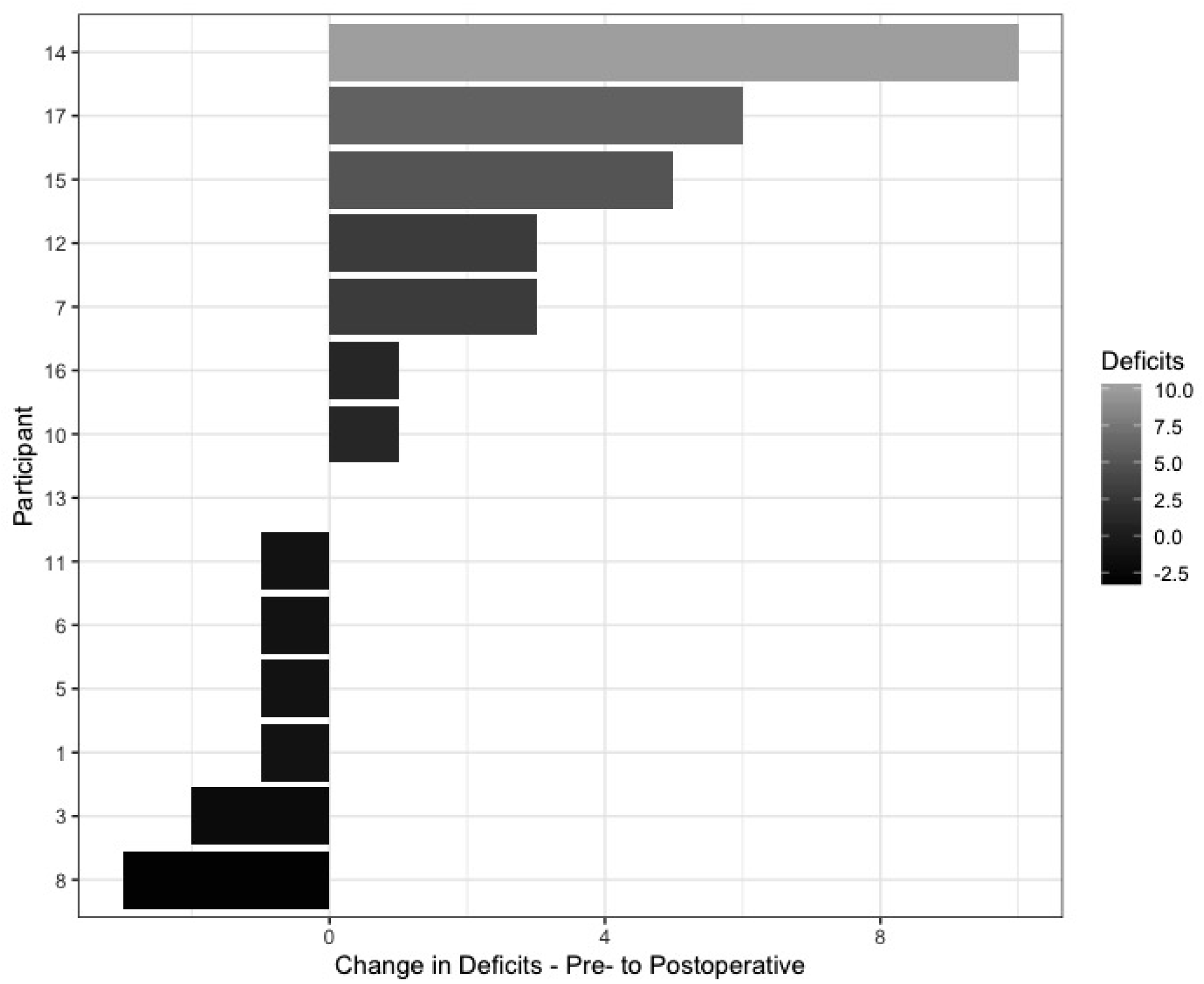
Change in the total number of deficits for each patient between pre- and postoperative traditional neuropsychology testing.

### OCS-BRIDGE Testing

As expected, OCS-bridge assessment required considerably less administration time (mean = 30.5 min, SD= 7.4) than traditional assessment (2-3 hours). Before surgery, 59%% (10/17) of participants had an impairment in at least one OCS-BRIDGE domain, with a mean of 0.94 impairments (SD = 1.08) per participant. Impairments before surgery occurred in the domains of numerical cognition (4), perception (3), attention (3), language (1), praxis (1), and verbal working memory (1). No participants showed an impairment in memory or prospective memory before surgery. Of the 39 individual cognitive tests included in the OCS-BRIDGE battery, 26 (66%) detected at least 1 impaired performance whilst performance on the remaining 13 was uniformly within the normal range.

### OCS-BRIDGE Longitudinal changes

OCS-BRIDGE’s relative brevity and ease of administration lent itself to longitudinal assessment as part of follow-up clinics.. The variation of postoperative impairments detected is shown in **Figure 2**. The greatest number were seen in attention and non-verbal skills. Of the 16 participants who had at least one post-operative assessment, 44% (7/16) had a reduced number of impairments by their last assessment, 25% (4/16) had the same, and 31% (5/16) showed an increase. Four participants who had multiple follow-ups showed a pattern of increased impairments on either post-operative or 3-month testing which resolved by their last follow-up date. Three of these four participants showed a transient increase of at least one impairment in perception, and two showed a transient increase in memory impairments. Ten of the sixteen participants who had at least one follow-up OCS-BRIDGE assessment ended the study with no impairment on formal testing.

**Figure 2.**
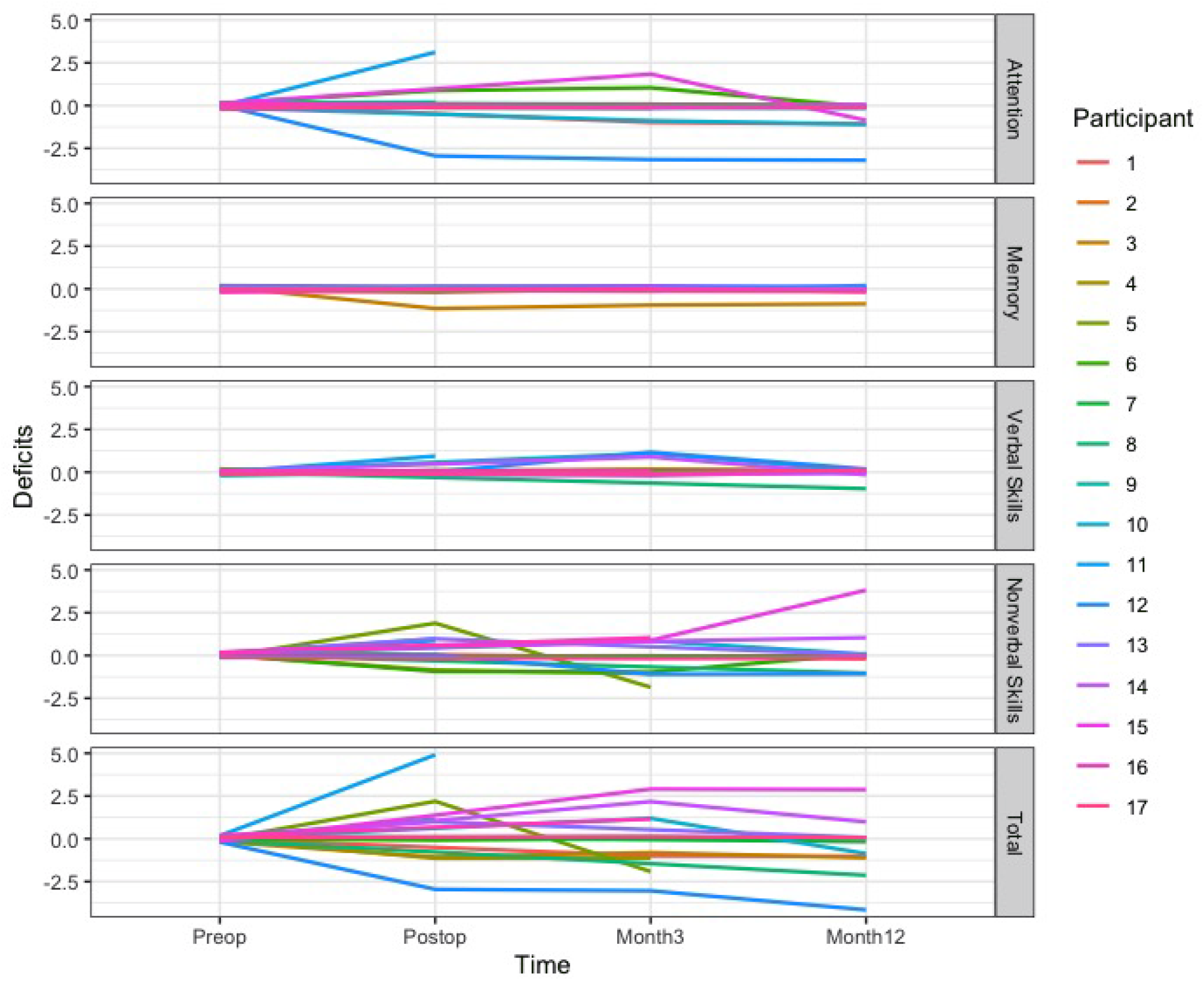
Number of cognitive deficits over time based on OCS-BRIDGE assessment preoperatively, postoperatively and at 3 and 12 month follow-ups.

### Comparison of OCS-BRIDGE with Traditional Neuropsychological Testing

As discussed, a proportion of the OCS-BRIDGE tests are extremely brief screening measures - the authors, therefore, applied a conservative strategy to avoid a high, and hence uninformative, level of false positives. It would therefore be predicted that OCS-BRIDGE would be less sensitive to impairments than in-depth traditional neuropsychological assessments. Figure 3A shows the total number of preoperative impairments detected in different domains using the two methods (**Figure 3A**). The tests in the two batteries vary quite widely but one area of commonality is in the forward and backward digit span tests. As shown in **Figure 3B**, the tests returned similar findings with no statistically significant differences on the forward (traditional mean = 6.29, SD = 1.40 vs. OCS-BRIDGE mean 6.76, SD = 0.90; t = -1.367, *P* = 0.1905) or backward tests (traditional mean = 4.47, SD = 1.28 vs. OCS-BRIDGE mean = 4.82, SD = 1.55; t= -0.972, *P*= 0.3456).

**Figure 3.**
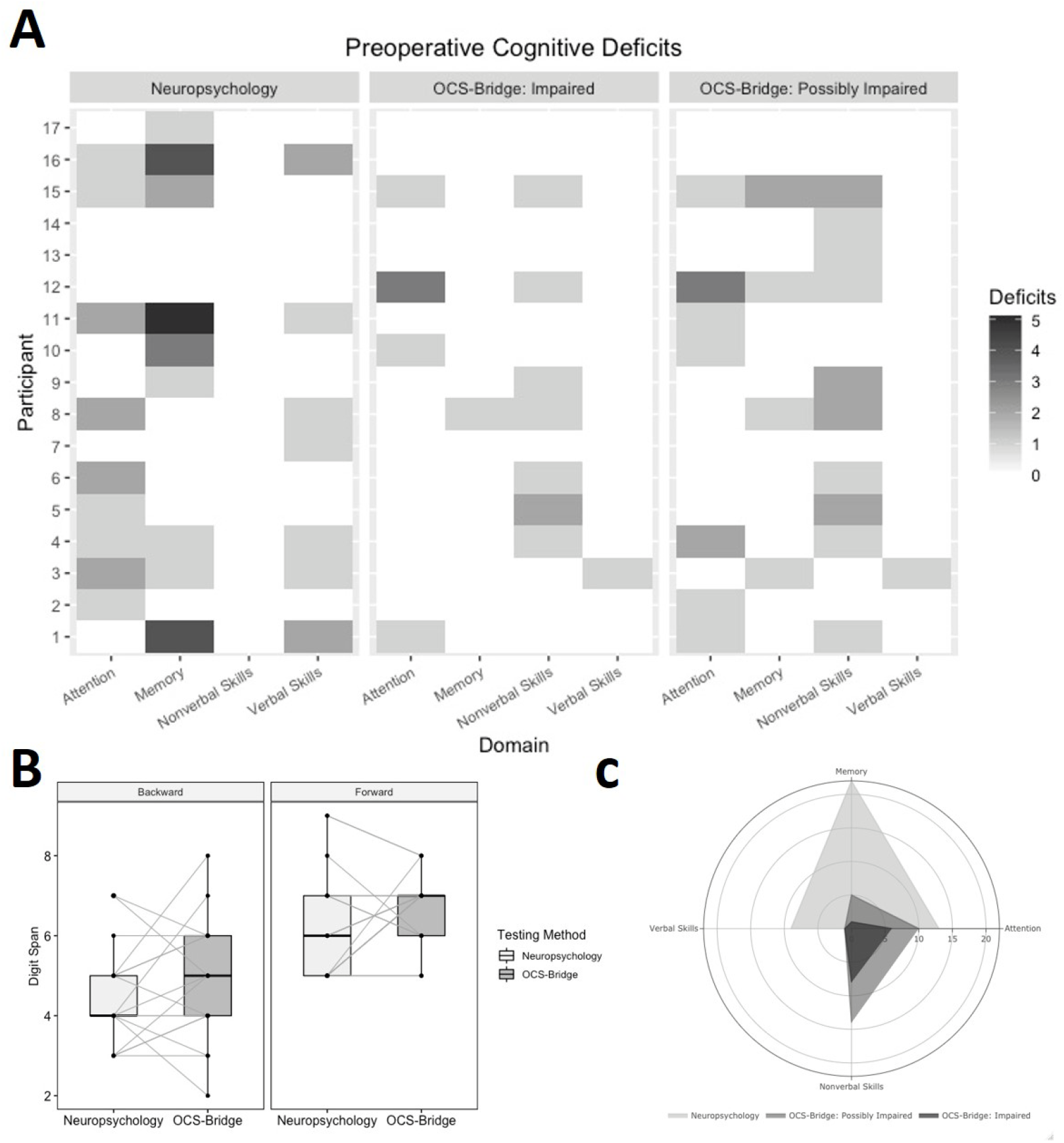
**A**. Cognitive deficits by domain from preoperative neuropsychological testing and OCS-BRIDGE assessments, **B**. Comparison of digit span analysis between neuropsychological testing and OCS-BRIDGE, C. Total preoperative deficits shown by neuropsychological testing and OCS-BRIDGE

Overall, traditional neuropsychological tests detected 44 preoperative impairments among the 17 participants in the four combined domains of attention, memory, verbal skills and non-verbal skills. OCS-BRIDGE detected 13 impairments and 28 possible impairments pre-operatively (**Figure 3C**). The average ratio of detected deficits was significantly higher for the neuropsychological assessments than for OCS-BRIDGE in the attention (preoperative, t=3.35, *P=*0.002; postoperative, t=2.64, *P=*0.016) and memory (preoperative, t=3.24, *P=*0.003; postoperative, t=1.79, *P=*0.088) domains. On the contrary, OCS-BRIDGE revealed a higher ratio of non-verbal skills deficits than neuropsychological assessments (preoperative, t=-3.11, *P=*0.004; postoperative, t=-3.16, *P=*0.005) (see **Table 2** for the average ratio of deficits per assessment and **Table S4** for the total number of deficits).

**Table 2.** Average ratio and standard deviation of cognitive deficits across patients for each domain and assessment. A positive *T* value indicates a higher ratio of deficits detected by traditional neuropsychology compared with OCS-BRIDGE.

|  |  | Neuropsychology<br>Average Ratio<br>(SD) | OCS-BRIDGE<br>Average Ratio (SD) | *T* value | *P* value |
| --- | --- | --- | --- | --- | --- |
| Attention | Preoperative | 0.255 (0.28) | 0.025 (0.56) | 3.35 | **0.002** |
|  | Postoperative | 0.262 (0.23) | 0.036 (0.76) | 2.64 | **0.016** |
| Memory | Preoperative | 0.078 (0.10) | 0 (0) | 3.24 | **0.003** |
|  | Postoperative | 0.131 (0.14) | 0.031 (0.09) | 1.79 | 0.088 |
| Non-verbal<br>skills | Preoperative | 0 (0) | 0.034 (0.04) | -3.11 | **0.004** |
|  | Postoperative | 0 (0) | 0.080 (0.10) | -3.16 | **0.005** |

Z-scores were calculated from both the neuropsychological testing and the OCS-BRIDGE data for each individual test **(Table 3)**. The only significant differences before and after surgery at item level (OCS-BRIDGE extinction.totalCorrect and boat.lineHeight; Neuropsychology AIMPB Story Immediate Recall and BMIPB Word list A6) did not survive correction for multiple comparisons. Item Z-scores were averaged over each domain, revealing a variety of shape distributions with a predominance of Normal (Neuropsychology verbal skills) and negatively skewed (memory, non-verbal skills, Executive Function) forms (**Figure S1**). No outliers were observed that may have overtly influenced statistical testing. Total and memory preoperative Z-score for neuropsychological testing were weakly correlated with OCS-BRIDGE Z-score across patients but significance did not survive multiple correction (Total, R=0.49, *P*_*uncorrected*_=0.045; Attention, R=0.27, *P*_*uncorrected*_=0.29; Memory, R=0.50, *P*_*uncorrected*_=0.041; Non Verbal Skills, R=0.06, *P*_*uncorrected*_=0.82; **Figure 4**). At the group level, only attention preoperative scores were significantly different between Neuropsychology and OCS-BRIDGE (*P*_*corrected*_=0.04. **Table 4**). To explore this further, unsupervised clustering was carried out to assess whether individual tests clustered into groups that were different from the original domains (**Figure 5)**. Clusters were named for their dominant test type. Cluster 1 included 10 tests, predominantly memory tests from the traditional battery, and included all but one test from AMIPB and BMIPB. Cluster 2 included 10 tests, 8 from the traditional battery and 2 from OCS-BRIDGE, and was mostly composed of Attention and Verbal Skills tests. Cluster 3 included only 4 tests, 3 of which were from the SALT attention task within the OCS-BRIDGE battery. Finally, Cluster 4 was the largest cluster, with 29 of the total tests, and included all but one of the tests of non-verbal skills, as well as 10 out of 14 of the OCS-BRIDGE attention tests. Tasks which included several individual tests, such as the OCS-BRIDGE Hearts and SALT tasks, clustered together, in Cluster 4 and Cluster 3 respectively. The Beck Depression Inventory fell into Cluster 1, while the Beck Anxiety Inventory fell into Cluster 4.

**Table 3.** Raw, standard deviation (SD) and Z-scored performance of each item included in traditional neuropsychology and OCS-BRIDGE. T-values and P-values result from a paired t-tests comparing scores before (preop) and after (postop) surgery.

| OCS-BRIDGE |  |  |  |  |  |  |  |
| --- | --- | --- | --- | --- | --- | --- | --- |
|  | Item name | Raw<sub>preop</sub> (SD) | Z<sub>preop</sub> | Raw<sub>postop</sub> (SD) | Z<sub>postop</sub> | t - value | P - value |
| Attention | hearts.overallScore | 29.31 (-1.08 ) | 0.34 | 28.63 (-1.41 ) | -0.06 | 1.33 | 0.20 |
|  | hearts.spaceAsymmetry | -0.002 (-0.03 ) | 0.03 | 0 (-0.07 ) | 0.10 | -0.19 | 0.85 |
|  | hearts.objectAsymmetry | 0.12 (-0.6 ) | 0.11 | 0.38 (-0.74 ) | 0.55 | -0.93 | 0.36 |
|  | hearts.perseverativeResponsesTo tal | 0 (0 ) | 0.06 | 0 (0 ) | 0.06 | 0.00 | 1.00 |
|  | hearts.organisationIndex | -2.64 (-0.76 ) | 0.16 | -2.13 (-0.63 ) | 0.70 | -1.64 | 0.11 |
|  | salt.overallFlashScore | 3.53 (-0.72 ) | -0.02 | 3.43 (-0.79 ) | -0.16 | 0.30 | 0.76 |
|  | salt.numberAllTargetsDetected | 35.71 (-0.69 ) | 0.18 | 35.29 (-1.5 ) | -0.28 | 0.96 | 0.35 |
|  | salt.RTCoefficient | -0.15 (-0.06 ) | 0.81 | -0.15 (-0.03 ) | 0.90 | -0.26 | 0.80 |
|  | salt.targetBias | 0 (-0.02 ) | -0.06 | 0 (-0.01 ) | 0.22 | -0.66 | 0.52 |
|  | salt.RTBias | 0.01 (-0.04 ) | 0.17 | 0.04 (-0.04 ) | 0.68 | -1.21 | 0.24 |
| Perception | extinction.totalCorrect | 17.94 (-0.24 ) | 0.12 | 17.43 (-0.79 ) | -1.30 | 2.48 | 0.02 |
|  | extinction.fieldProblem | 0 (0 ) | -0.02 | -0.14 (-0.38 ) | -0.61 | 1.61 | 0.12 |
|  | extinction.leftExtinction | 0 (0 ) | -0.07 | 0 (0 ) | -0.07 | 0.00 | 1.00 |
|  | extinction.rightExtinction | 0 (0 ) | -0.08 | 0.14 (-0.9 ) | 0.34 | -0.68 | 0.51 |
|  | boat.lineHeight | -0.99 (-0.23 ) | -0.78 | -1.26 (-0.23 ) | -2.21 | 2.70 | 0.01 |
|  | boat.meanDeviation | -0.56 (-2.29 ) | -0.11 | -0.4 (-1.4 ) | -0.05 | -0.18 | 0.86 |
|  | pstl.overallScore | 6 (0 ) | 0.13 | 6 (0 ) | 0.13 | 0.00 | 1.00 |
| Memory | memory.verbalFreeRecall | 3.06 (-0.97 ) | -0.18 | 3 (-1.51 ) | -0.25 | 0.12 | 0.91 |
|  | memory.verbalOverallRecall | 3.88 (-0.33 ) | 0.02 | 3.75 (-0.46 ) | -0.34 | 0.82 | 0.42 |
|  | memory.episodicMemoryScore | 4 (0 ) | 0.18 | 4 (0 ) | 0.18 | 0.00 | 1.00 |
|  | orientation.overallScore | 4 (0 ) | 0.17 | 3.88 (-0.35 ) | -0.61 | 1.49 | 0.15 |
|  | fins.forwardDigitSpan | 6.76 (-0.9 ) | -0.20 | 6.14 (-1.07 ) | -0.66 | 1.46 | 0.16 |
|  | fins.backwardDigitSpan | 4.82 (-1.55 ) | -0.39 | 4.43 (-1.4 ) | -0.65 | 0.58 | 0.57 |
|  | pstp.prospectiveScore | 4 (0 ) | 0.71 | 4 (0 ) | 0.71 | 0.00 | 1.00 |
|  | pstp.retrospectiveScore | 4 (0 ) | 0.68 | 4 (0 ) | 0.68 | 0.00 | 1.00 |
| Language | Naming.overallScore | 4 (0 ) | 0.32 | 4 (0 ) | 0.32 | 0.00 | 1.00 |
|  | semantics.overallScore | 3 (0 ) | 0.06 | 3 (0 ) | 0.06 | 0.00 | 1.00 |
|  | reading.overallScore | 15 (0 ) | 0.34 | 15 (0 ) | 0.34 | 0.00 | 1.00 |
| Praxis | imitation.handScore | 5.88 (-0.49 ) | 0.00 | 5.88 (-0.35 ) | -0.02 | 0.04 | 0.97 |
|  | imitation.fingerScore | 6 (0 ) | 0.20 | 6 (0 ) | 0.20 | 0.00 | 1.00 |
|  | imitation.overallScore | 11.88 (-0.49 ) | 0.12 | 11.88 (-0.35 ) | 0.11 | 0.04 | 0.97 |
| Number | numerical.overallArithmeticScore | 3.47 (-0.94 ) | -0.87 | 3.63 (-0.52 ) | -0.50 | -0.43 | 0.67 |
|  | numerical.overallWritingScore | 2.88 (-0.49 ) | -1.00 | 2.88 (-0.35 ) | -1.07 | 0.04 | 0.97 |
| Neuropsychological Assessments |  |  |  |  |  |  |  |
|  | Item name | Raw<sub>preop</sub> (SD) | Z<sub>preop</sub> | Raw<sub>postop</sub> (SD) | Z<sub>postop</sub> | t - value | P - value |
| Attention | WAIS-IV.Digit.Span.Forward | 9.88 (-2.5 ) | -0.04 | 9.5 (-2.24 ) | -0.17 | 0.44 | 0.66 |
|  | WAIS-IV.Digit.Span.Backward | 8.12 (-2.2 ) | -0.63 | 7.21 (-1.85 ) | -0.93 | 1.22 | 0.23 |
|  | WAIS-IV.Digit.Symbol | 66.29 (-13.85 ) | -0.18 | 66.07 (-15.43 ) | -0.19 | 0.04 | 0.97 |
| Non-Verbal Skills | BMIPB.Complex.Figure.copy | 79.12 (-1.17 ) | 0.30 | 79.38 (-0.87 ) | 0.40 | -0.69 | 0.50 |
|  | VOSP.Object.Decision | 18.29 (-1.16 ) | -0.19 | 19 (-1.22 ) | 0.25 | -1.61 | 0.12 |
|  | VOSP.Number.Location | 9.41 (-0.87 ) | 0.01 | 9.38 (-0.65 ) | -0.01 | 0.09 | 0.93 |
|  | VOSP.Cube.Analysis | 9.76 (-0.56 ) | 0.39 | 9.77 (-0.44 ) | 0.39 | -0.02 | 0.98 |
| Memory | AMIPB.Story-Immediate.Recall | 34.71 (-8.99 ) | 0.17 | 27 (-9.04 ) | -0.59 | 2.37 | 0.02 |
|  | AMIPB.Story-Delayed.Recall | 32.82 (-11.14 ) | 0.18 | 27.54 (-11.4 ) | -0.32 | 1.28 | 0.21 |
|  | BMIPB.Word.List.A1-A15 | 54.59 (-9.1 ) | -0.22 | 50.14 (-9.36 ) | -0.84 | 1.34 | 0.19 |
|  | BMIPB.Word.List.A6 | 12.82 (-2.53 ) | 0.29 | 10.5 (-2.95 ) | -0.64 | 2.36 | 0.03 |
|  | BMIPB.Word.List Word.Recognition | 28.71 (-0.99 ) | 0.08 | 27.64 (-2.47 ) | -0.74 | 1.63 | 0.11 |
|  | BMIPB.Word.List List.Recognition | 28.94 (-1.56 ) | -0.04 | 27.5 (-3.61 ) | -1.00 | 1.49 | 0.15 |
|  | BMIPB.Complex.Figure Immediate.Recall | 65.47 (-7.8 ) | 0.02 | 61.36 (-13 ) | -0.39 | 1.09 | 0.28 |
|  | BMIPB.Complex.Figure Delayed.Recall. | 63.24 (-8.21 ) | 0.11 | 59.57 (-13.36 ) | -0.23 | 0.94 | 0.36 |
|  | BMIPB.Design.Learning-A1-A15 | 37.35 (-6.19 ) | -0.01 | 35.14 (-7.22 ) | -0.38 | 0.92 | 0.37 |
|  | BMIPB.Design.Learning-A6 | 8.29 (-1.16 ) | 0.07 | 8.14 (-1.61 ) | -0.04 | 0.30 | 0.76 |
|  | BMIPB.Design.Learning Recognition | 39.41 (-1.18 ) | 0.32 | 38.43 (-2.59 ) | -0.12 | 1.40 | 0.17 |
|  | BMIPB.Design.Learning Identification | 9.59 (-1 ) | 0.13 | 9 (-1.62 ) | -0.27 | 1.24 | 0.22 |
| Language | Letter.Fluency | 37.82 (-14.55 ) | -0.25 | 30.71 (-15.04 ) | -0.91 | 1.33 | 0.19 |
|  | Semantic.Fluency | 20.71 (-5.92 ) | 0.22 | 16.5 (-5.68 ) | -0.79 | 2.00 | 0.05 |
|  | Graded.Naming.Test | 19.65 (-3.87 ) | -0.66 | 19.57 (-5 ) | -0.68 | 0.05 | 0.96 |
|  | Syntactic.Speech.Comprehension | 24.88 (-0.93 ) | -0.08 | 24.57 (-1.99 ) | -0.29 | 0.57 | 0.57 |
| Executive Functioning | Hayling.Initiation.(time) | -3.88 (-3.76 ) | -0.76 | -2.5 (-1.61 ) | -0.13 | -1.28 | 0.21 |
|  | Hayling.Inhibition.(time) | -21.24 (-16.06 ) | -0.36 | -23 (-21.01 ) | -0.49 | 0.27 | 0.79 |
|  | Hayling.Inhibition.(score) | 10.94 (-3.23 ) | -0.04 | 10.86 (-3.72 ) | -0.07 | 0.07 | 0.95 |
|  | Brixton | -10.94 (-4.49 ) | 0.89 | -10.92 (-7.1 ) | 0.89 | -0.01 | 0.99 |

**Table 4.**
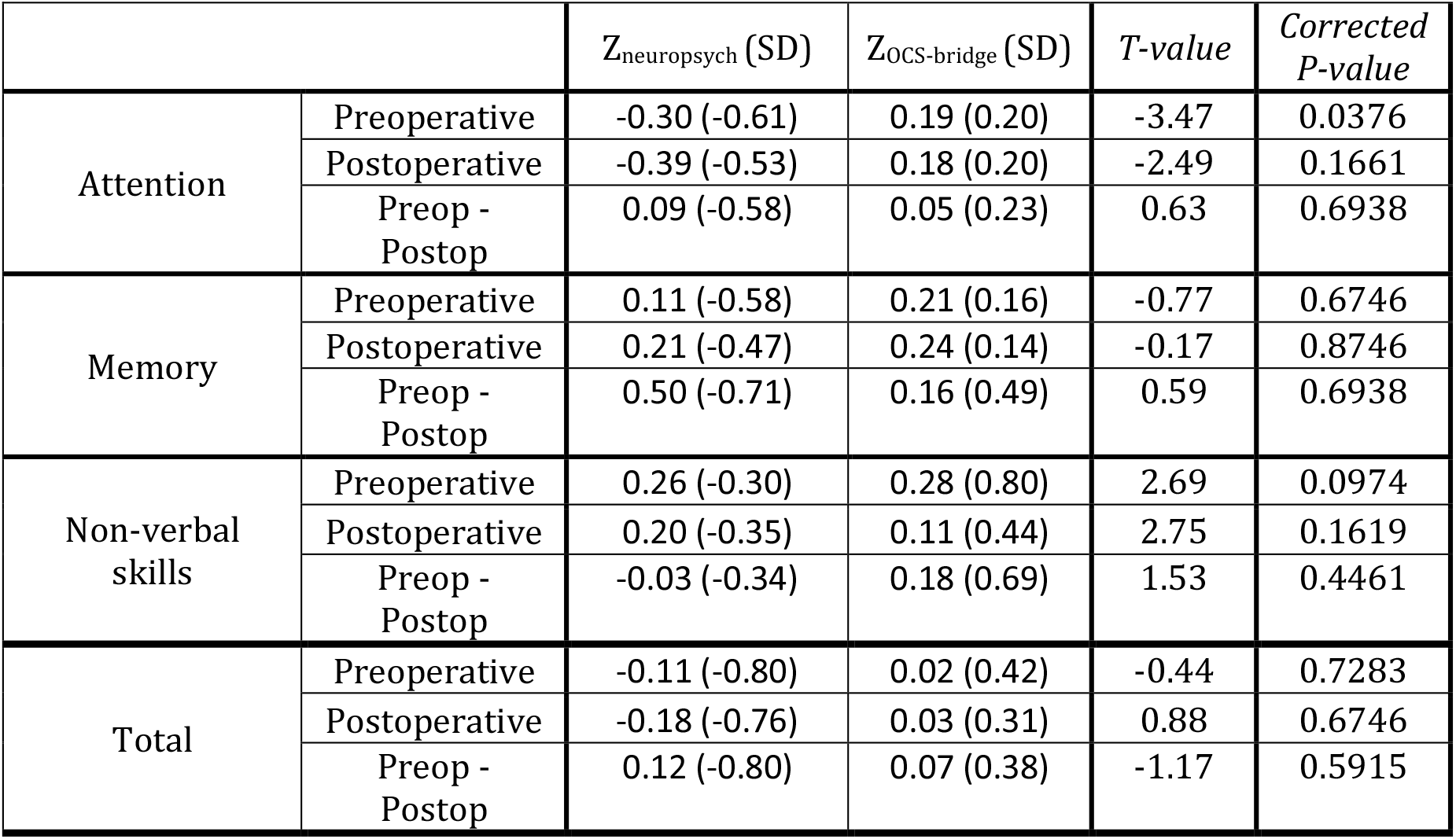
Differences in cognitive performance between traditional neuropsychology and OCS-BRIDGE. Z_neuropsych_ and Z_OCS-bridge_ indicate the average Z-score cognitive performance and standard deviation (SD) of each domain assessed by neuropsychological testing and OCS-bridge screening respectively.

**Figure 4.**
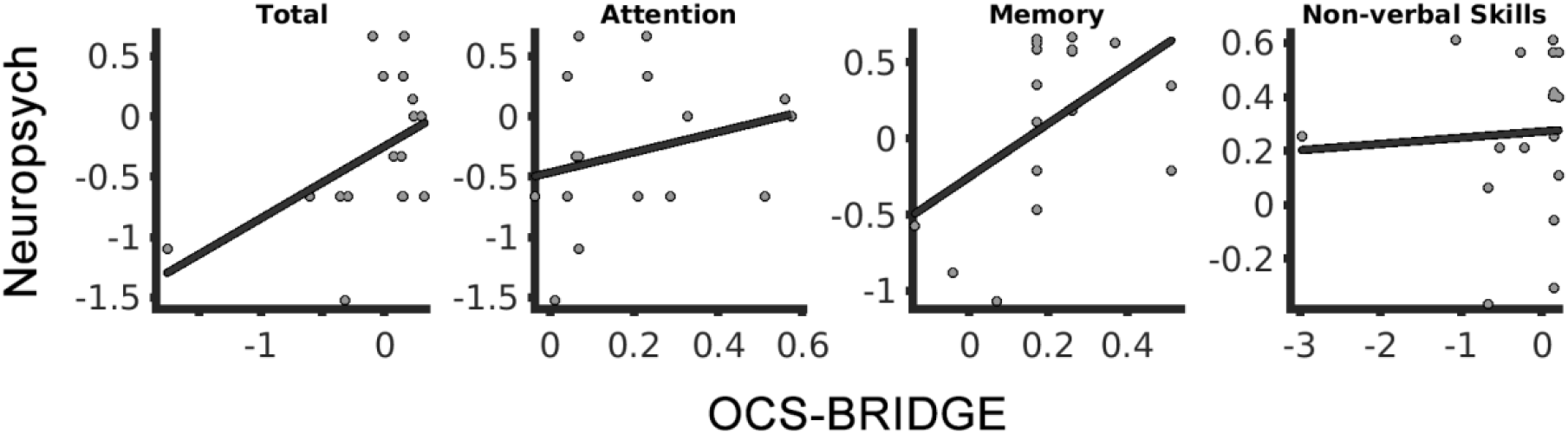
Association between Z-scores derived from OCS-BRIDGE and neuropsychological testing. Plots for overlapping domains (Attention, Memory and Non-verbal skills) and for the total average across domains (Total).

**Figure 5.**
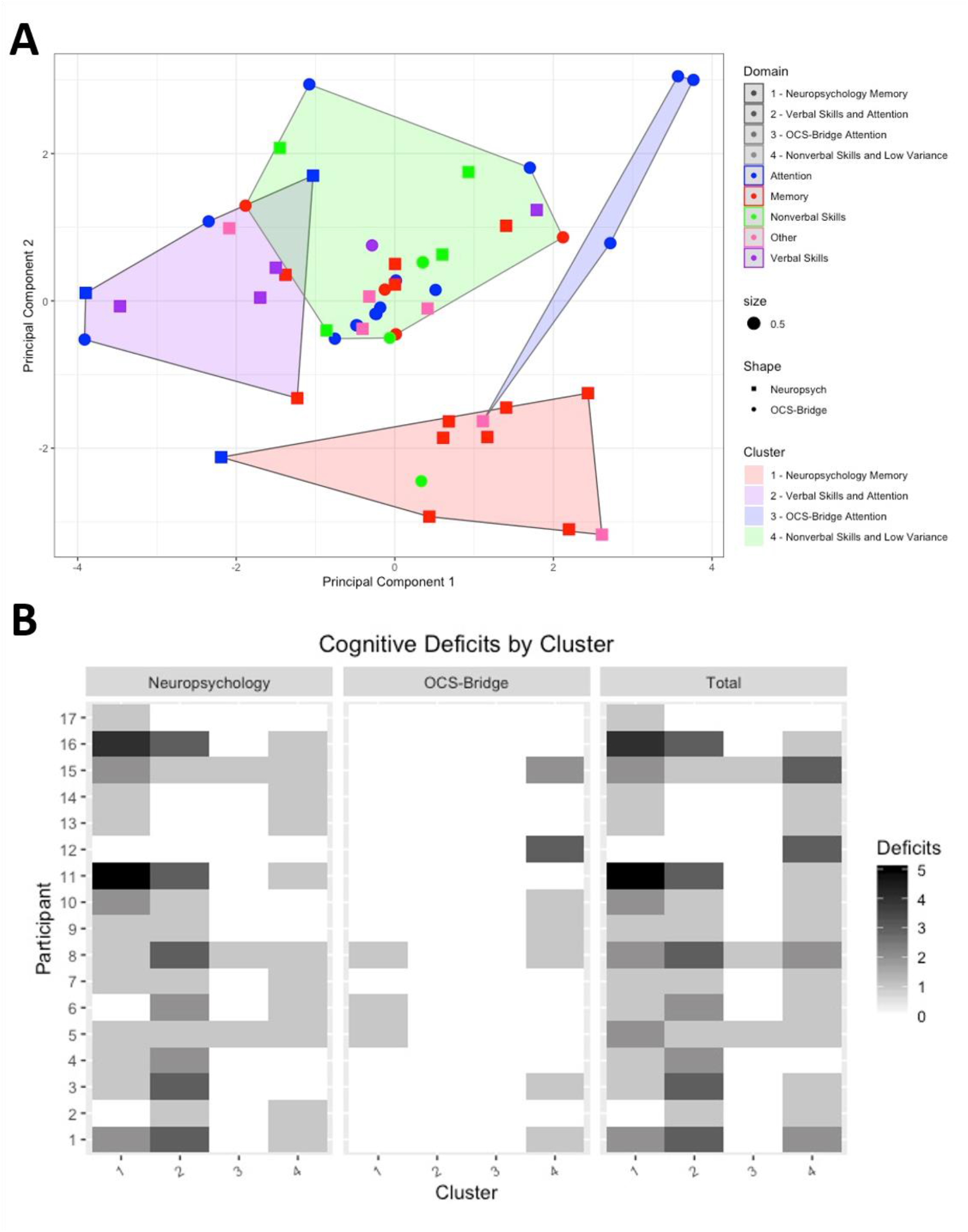
K-means clustering of individual cognitive tests and validation against deficit numbers. **A**. Principal components analysis plotted versus the k-means clustering results, **B**. Cognitive deficits for each participant by cluster, both before and after surgery.

## DISCUSSION

Diffuse glioma can cause cognitive impairments and identifying these is important in informing patients and families, in improving management, and in evaluating outcomes. Traditional neuropsychological testing is resource-intensive, making it difficult to perform at multiple time points during the patient journey (Taphoorn & Klein, 2004). Tablet-based assessments tools such as OCS-BRIDGE provide rapid, easy-to-use alternatives to traditional pen-and-paper testing, yet little work has been done to demonstrate the validity of these tests in this population, or to assess their relative strengths and weaknesses when compared with traditional batteries. This paper presents early data from a cohort of 17 participants with diffuse glioma who underwent traditional and OCS-BRIDGE assessment, both before and after surgery, and at several follow-ups. The results show that both methods detected cognitive impairments in participants who did not primarily present with self-reported cognitive problems

Results revealed that, when compared with a 2-3 hour traditional assessment, a 30-min OCS-BRIDGE assessment significantly detected fewer impairments in the areas of attention and memory. However, OCS-BRIDGE significantly detected more impairments in non-verbal skills. Memory and attention screening using only a few items cannot make the kind of fine distinctions possible with a longer test, and as a consequence can have a more conservative threshold for impairment. In contrast, OCS-BRIDGE measures with reaction time assessments can make very fine individual difference distinctions. The results suggest that a combined approach, using traditional assessment in those areas where brief screening may be less sensitive, and OCS-BRIDGE style measures for reaction time and perceptual tasks, may be most effective.

Comparisons based on Z-scores showed no significant differences between preoperative and postoperative scores at the individual item level, revealing that single items have insufficient statistical power for detecting average decreases in performance even for individuals undergoing major surgery. However, the lack of significance may be driven by our limited sample size or by the challenges of correcting for multiple comparisons when a large number of tests are considered. Additionally, by grouping items into larger domains, we were able to compare assessment methods. The positive, but nonsignificant after correction, correlation between OCS-BRIDGE and Neuropsychological assessment suggest an effect that may be confirmed in larger samples. Nevertheless, the low explained variance is undoubtedly reflecting some degree of disagreement that in turn suggests a lower power for group-based Z-scores that compare assessments based on means and standard deviations obtained from patients. This could be particularly challenging for a brain tumour population with highly heterogeneous cognitive profiles. As this Z-scored analysis retains the variability of scores even for patients’ performances that are within normative ranges (i.e., an unthresholded approach), it may have masked the differences in low performance domains shown by the analysis based on the number of detected deficits, as discussed above. Thought needs to be given to the practicalities, duration and tolerability of assessment as well as the sensitivity of the measures in finding the most appropriate balance for particular clinical groups or individuals.

### Advantages of OCS-BRIDGE

There are several advantages of using a tablet-based system in conjunction with traditional neuropsychological testing in glioma patients. OCS-BRIDGE is easy to transport and can be administered anywhere that is relatively quiet and free from distraction. It is easy to administer in a standardized way without the necessity of a highly trained neuropsychologist at the point of administration. It automatically captures patient responses and scores and reports on performance saving time and reducing errors. Ease of use also presents the possibility that these assessments could be modified to be performed by the patient independently, furthering their potential for tracking subtle changes in cognitive functioning over time during recovery. As is observed here, an app-based system may pick up subtle differences in certain areas of performance missed by traditional testing, such as precise reaction times and accurate visual acuity scores.

### Complementary Role of Traditional and App-based Assessment

Cluster analyses have been extensively exploited in neuropsychology to better understand the relationship between test scores and for patient classification (Benassi et al., 2020; Morris et al., 1981). Here, we use this methodology to show that items are grouped according not only to a domain criterion but also following an assessment criterion (traditional vs. App-based). The latter is not surprising given that traditional neuropsychological testing exhibited higher sensitivity to impairments in memory. There are no inherent barriers to recreating in-depth memory assessments in app form, as is illustrated by the digit span measures that, in both cases here, began at a very easy level and continued until a participant consistently failed. The clustering also identified a subset of items with very limited sensitivity for detecting impairments (Cluster 3), suggesting that this could be potentially exploited for discriminating relevant assessments. The OCS memory measures were primarily developed for extremely fast screening in an acute stroke setting and may indeed be less suitable for the diffuse glioma population. In contrast, a tablet’s ability to capture reaction time with millisecond precision gives OCS-BRIDGE some sensitivity advantages over traditional neuropsychological assessment. The current results would certainly suggest that combining the strengths of app and traditional approaches would be warranted in the diffuse glioma population (or that more suitable app measures of memory are used). An issue here is whether to tailor a diffuse glioma battery to the time when they may be most fatigued/intolerant of a long testing session (post-surgery) or to shorten/lengthen testing as appropriate for each stage.

Quite a few measures in the two batteries used here had ceiling effects, that is, no participant scored less than perfectly. The reason for their inclusion was that there are important neuropsychological conditions to which such tests are sensitive that may have arisen in a larger sample. In selecting tests for a particular clinical population, the risks of failing to detect an impairment must be weighed against the costs and time of giving that assessment. In that respect, studies such as this are useful for informing such decisions.

Another factor supporting a complementary is the importance of the neuropsychologist, rather than the particular tests used. As discussed, uninterpreted test results can encourage a binary view that a function is either impaired or unimpaired. In contrast, it is possible that an individual who has fallen from the 95th to 15th percentile on a particular test has likely suffered a significant impairment regardless of a test finding performance to be in the ‘normal range’. Neuropsychologists use interviews about educational and occupational history, the experiences of patients and family members and the patterns across tests to think about what scores are likely to mean and the implications for management, rehabilitation and so on. There is currently no app for that. It is important that staff using app-based assessments have appropriate training in what the results do, and do not, mean.

The results presented above indicate potentially complementary roles of different modalities of cognitive testing in the assessment and postoperative follow-up of glioma patients. Traditional testing administered in a clinical setting by a qualified neuropsychologist demonstrated high sensitivity to cognitive impairments, particularly in the domains of memory and verbal skills, when compared with the OCS-BRIDGE tablet-based system. The tablet-based system showed higher sensitivity to impairments in non-verbal skills including impairments in perception, and took less time, expertise, and resources to administer and analyze. These differences indicate that there may be a role for a combined assessment schedule using both modalities for glioma patients in future to gain a more comprehensive understanding of their functional abilities pre- and post-operatively and as they recover in the longer term.

Combining cognitive assessments in an optimal way represents a major challenge. Existing literature has demonstrated that multiple tests used together can facilitate the diagnosis of neurodevelopmental conditions (Doyle et al., 2000; Perugini et al., 2000). However, the criteria for distinguishing clinical differences through battery test combination remains an open question. A liberal threshold considering deficits as those domains showing just at least one impairment in a test would have higher sensitivity and worse specificity than more conservative thresholds. Notwithstanding, (Lovejoy et al., 1999) showed that a liberal threshold of deficits in at least one out of six tests as signifying impairment provides the best overall classification rates of ADHD (*i*.*e*. the improvement in specificity does not outweigh the decrease in sensitivity). This is also of major relevance in our cohort of brain tumour patients where sensitivity is more critical than specificity (as patients with nonidentified deficits -False Negatives-will not receive appropriate training or treatment while the contrary does not risk significant harm to the patient). Although systematic comparisons will be necessary to optimally combine traditional neuropsychological and app-based assessments, the combination will need to be tailored to each neuropsychiatric condition and should balance sensitivity and specificity according to the impact that false positives and false negatives have on patients.

### Study limitations and constraints on generality

Beyond the reduced sample size, attrition represents an additional limitation of this study. In particular, the immediate post-operative assessment was often limited by fatigue, while follow-up data was on occasion precluded by the participants’ geographical distance from centralized assessment. In a related manner, there were logistical and technical limitations to the assessments, which is not entirely unexpected during the beta-testing phase and with such an ambitious study protocol.

All patients had the pre-operative imaging appearances of a diffuse glioma (non-enhancing and without oedema or mass effect), however subsequent pathological examination revealed a range of histological diagnoses (**Table 1**). There were other sources of heterogeneity such as tumour location, tumour size and treatment - some participants received only surgical intervention whilst others received follow-up chemotherapy or radiotherapy, which is itself associated with cognitive impairment risk (Lawrie et al., 2019; Wefel et al., 2011, 2016). Unfortunately, it is impossible to comment on the differential contributions of treatment regimens to cognitive functioning at this sample size. Nevertheless, these data present a ‘real-world’ view of treatment pathways that will be better reflective of outcomes encountered in series outside of clinical research studies. Finally, there are inevitable issues surrounding practice effects when using the same cognitive assessment tools over time in the same population (Barron et al., 2015; Cacciamani et al., 2018). A balance is required between the direct comparability of assessments at different time points and the potential for practice effects.

## Conclusions

Cognitive function in patients with diffuse glioma during early treatment is a complex and dynamic interplay between multiple factors. Both traditional and app-based assessments have complementary roles in understanding cognitive function. This work provides the framework for robust, objective, and accessible assessment across multiple centres. Such data will be useful not only clinically at the individual level, but also for generating the necessary large datasets required to better understand cognitive outcomes in general.

## Data Availability

In accordance with ethics requirements, data will be made available to collaborating centres upon reasonable request.

## Acknowledgements

We thank all patients for their generous involvement in the study. We also thank Luca Villa, Rohit Sinha and Jessica Ingham for their contribution to the study. RRG is funded by a Guarantors of Brain Post-Doctoral Fellowship award, by a Cancer Research UK Cambridge Centre RG86786 (CRUK grant ref: A25117). grant and by the EMERGIA Junta de Andalucía program. YE is funded by a Royal Society Dorothy Hodgkin Research Fellowship. MA is funded by the Cambridge Trust – Yousef Jameel Scholarship. SJP is supported by the National Institute for Health Career Development Fellowship (CDF-2018-11-ST2-003). This report is independent research supported by the National Institute of Research (NIHR Career Development Fellowship, Mr Stephen Price, CDF-2018-11-ST2-003). The views expressed in this publication are those of the authors and not necessarily those of the NHS, the National Institute for Health Research or the Department of Health and Social Care. MGH received an award from The Brain Tumour Charity (ref: RG86218) to fund this work.

## Conflict of Interest/Competing interests

There are no known conflicts of interest for any author of this manuscript

## Code availability

Code will be made available upon request.

## Notes

### Competing Interest Statement

The authors have declared no competing interest.

### Funding Statement

RRG is funded by a Guarantors of Brain PostDoctoral Fellowship award, by a Cancer Research UK Cambridge Centre RG86786 grant (CRUK grant ref: A25117) and by the EMERGIA Junta de Andalucia program. YE is funded by a Royal Society Dorothy Hodgkin Research Fellowship. MA is funded by the Cambridge Trust Yousef Jameel Scholarship. SJP is supported by the National Institute for Health Career Development Fellowship (CDF 2018 11 ST2 003). This report is independent research supported by the National Institute of Research (NIHR Career Development Fellowship, Mr Stephen Price, CDF-2018-11-ST2-003).

### Author Declarations

This study was approved by the Cambridge Central Research Ethics Committee (protocol number 16/EE/0151).

