## Supplemental Information for "Assessment of neuropsychological function in brain tumour treatment: A comparison of traditional neuropsychological assessment with app-based cognitive screening"

### Supplementary material

Rafael Romero-Garcia[1*], Mallory Owen[1*], Alexa McDonald[2], Emma Woodberry[2], Moataz Assem[3], Pedro Coelho[4], Rob C Morris[5], Stephen J Price[5], Tom Santarius[5,6], John Suckling[1,7,8], Tom Manly[8], Yaara Erez[3], Michael G Hart[1]

*****These authors contributed equally

1. University of Cambridge School of Clinical Medicine, 2. Department of Neuropsychology, Cambridge University Hospitals NHS Foundation Trust, 3. MRC Cognition and Brain Sciences Unit, University of Cambridge, 4. Neurophys Limited, 5. Department of Neurosurgery, Addenbrooke's hospital, Cambridge, 6. Physiology, Development and Neuroscience, University of Cambridge, 7. Behavioural and Clinical Neuroscience Institute, University of Cambridge; 8. Cambridge and Peterborough NHS Foundation Trust.

**Figure S1.** Distribution of z-scores for individual items across patients grouped by major domains (i.e., each count represents the score of one item in one of the patients' assessment) Values less than 0 indicate items where brain tumour patients showed lower performance than healthy controls.

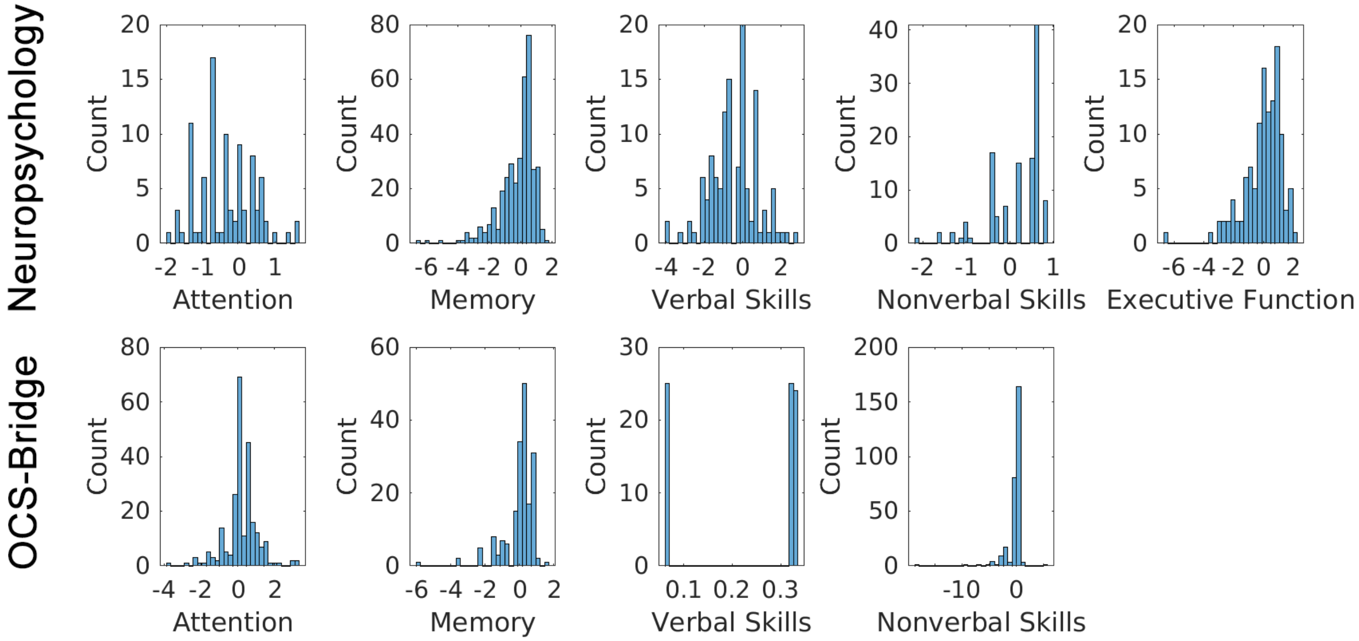

**Table S1**. Individual cognitive tests included in the OCS-Bridge assessment and the traditional neuropsychology battery

| **OCS-Bridge Assessment** | |
|---|---|
| **Oxford Cognitive Assessment** | |
| **Domain** | **Test** |
| Attention | Hearts Accuracy |
| Attention | Space Neglect |
| Attention | Space Neglect |
| Attention | Object Neglect |
| Attention | Object Neglect |
| Attention | Perseveration |
| Attention | Hearts Organization Index |
| Perception | Visual PerceptionTotal |
| Perception | Visual Field |
| Perception | Visual Field |
| Perception | Visual Extinction Left |
| Perception | Visual Extinction Right |
| Memory | Memory Free Verbal |
| Memory | Overall Verbal Memory |
| Memory | Episodic Memory |
| Memory | Orientation |
| Language | Picture Naming |
| Language | Semantics |
| Language | Sentence Reading Words Recalled |
| Praxis | Hand Positions |
| Praxis | Finger Positions |
| Praxis | Overall |
| Number | Calculation |
| Number | Number Writing |
| **Cambridge Assessment** | |
| Perception | Boat Visual Acuity |
| Perception | Line bisection |
| Perception | Line bisection |
| Perception | Postcards object perception |
| Attention | SALT sustained attention |
| Attention | Target detection |
| Attention | Consistency of Attention |
| Attention | Target space bias |
| Attention | Target space bias |
| Attention | Reaction time space bias |
| Attention | Reaction time space bias |
| Verbal Working Memory | Forward Digit Span |
| Verbal Working Memory | Backwards Digit Span |

| **Neuropsychology Battery** | |
|---|---|
| **Domain** | **Test** |
| Verbal Memory | AMIPB Story – Immediate Recall |
| Verbal Memory | AMIPB Story – Delayed Recall |
| Verbal Memory | BMIPB Word List - A1-A5 |
| Verbal Memory | BMIPB Word List - A6 |
| Verbal Memory | BMIPB Word List - Word recognition |
| Verbal Memory | BMIPB Word List - List recognition |
| Nonverbal Memory | BMIPB Complex Figure – Immediate Recall |
| Nonverbal Memory | BMIPB Complex Figure – Delayed Recall |
| Nonverbal Memory | BMIPB Design Learning - A1-A5 |
| Nonverbal Memory | BMIPB Design Learning - A6 |
| Nonverbal Memory | BMIPB Design Learning - Design Recognition |
| Nonverbal Memory | BMIPB Design Learning - Design Identification |
| Verbal Skills | Test of Premorbid Functioning |
| Verbal Skills | Letter Fluency – FAS |
| Verbal Skills | Semantic Fluency – Animals |
| Verbal Skills | Graded Naming Test |
| Verbal Skills | Syntactic Speech Comprehension |
| Nonverbal skills | BMIPB Complex Figure copy |
| Nonverbal skills | VOSP Object Decision |
| Nonverbal skills | VOSP Number Location |
| Nonverbal skills | VOSP Cube Analysis |
| Attention | WAIS-IV Digit Span - Forward |
| Attention | WAIS-IV Digit Span - Backward |
| Executive Function | Initiation - Time |
| Executive Function | Inhibition - Time |
| Executive Function | Inhibition - Score |
| Executive Function | Brixton |
| Mood | Beck Anxiety Inventory |
| Mood | Beck Depression Inventory |

| | |
|---|---|
| Prospective Memory | Prospective Memory |
| Prospective Memory | Retrospective Memory |

**Table S2.** Combined domains across neuropsychological testing and OCS-Bridge testing.

| **Combined Domain** | **Neuropsychology** | **OCS-Bridge** |
|---|---|---|
| Attention | Attention | Attention |
| Memory | Nonverbal Memory<br>Verbal Memory | Memory<br>Prospective Memory<br>Verbal Working Memory |
| Verbal Skills | Verbal Skills | Language |
| Nonverbal Skills | Nonverbal Skills | Praxis<br>Number<br>Perception |
| Executive Function | Executive Function | - |
| Mood | Mood | - |

**Table S3.** OCS-Bridge screening and neuropsychological assessments completed by each participant.

| ID | Pre-op | | Post-op | | Month 3 | Month 12 |
|---|---|---|---|---|---|---|
| | OCS-Bridge | Psychology | OCS-Bridge | Psychology | OCS-Bridge | OCS-Bridge |
| 1 | Done | Done | Not tolerated | Done | Done | Done |
| 2 | Done | Done | Done | Done | Done | Done |
| 3 | Done | Done | Not tolerated | Not available | Failed | Failed |
| 4 | Done | Done | Done | Done | Done | Logistics |
| 5 | Done | Done | Done | Done | Done | Failed |
| 6 | Done | Done | Done | Done | Done | Done |
| 7 | Done | Done | Not tolerated | Done | Failed | Done |
| 8 | Done | Done | Failed | Done | Failed | Done |
| 9 | Done | Done | Done | Failed | Failed | Failed |
| 10 | Done | Done | Done | Done | Failed | Failed |
| 11 | Done | Done | Failed | Done | Done | Done |
| 12 | Done | Done | Done | Done | Done | Done |
| 13 | Done | Done | Done | Done | Logistics | Done |
| 14 | Done | Done | Dysphonic | Done | Done | Done |
| 15 | Done | Done | Logistical | Done | Done | Done |
| 16 | Done | Done | MA syndrom | Done | Done | Failed |
| 17 | Done | Done | Dysphasic | Done | Done | Done |
| Totals | 17/17 | 17/17 | 8/17 | 15/17 | 10/17 | 11/17 |

**Table S4.** Average total number of cognitive deficits and standard deviations across patients for each domain and assessment. A positive T value indicates a larger number of deficits detected by traditional neuropsychology compared with OCS-BRIDGE.

| | | Neuropsychology Average (SD) | OCS-BRIDGE Average (SD) | *T* value | *P* value |
|---|---|---|---|---|---|
| Attention | Preoperative | 0.765 (0.83) | 0.353 (0.79) | 1.48 | 0.15 |
| | Postoperative | 0.786 (0.70) | 0.500 (1.07) | 0.76 | 0.46 |
| Memory | Preoperative | 0.941 (1.20) | 0 (0) | 3.24 | **0.003** |
| | Postoperative | 1.571 (1.70) | 0.125 (0.35) | 2.36 | **0.03** |
| Non-verbal skills | Preoperative | 0 (0) | 0.471 (0.62) | -3.11 | **0.004** |
| | Postoperative | 0 (0) | 1.125 (1.36) | -3.16 | **0.005** |
